# Omicron-associated mortality for principal causes other than COVID-19, including mortality with a confirmed SARS-CoV-2 infection, and ICU admissions with an Omicron infection in adults aged over 60 years in France

**DOI:** 10.1101/2022.12.15.22283529

**Authors:** Edward Goldstein

**Affiliations:** Massachusetts Eye and Ear, Harvard Medical School, Boston MA, USA

## Abstract

**Background:** With the emergence of the Omicron variant, an increasing proportion of SARS-CoV-2 associated deaths have a principal cause of death other than COVID-19. In France, between Nov. 1, 2021 --July 31, 2022, in addition to 33,353 deaths with the principal cause of COVID-19, there were 9,638 deaths with a confirmed SARS-CoV-2 infection with a principal cause of death other than COVID-19 (as well as SARS-CoV-2-associated deaths with an undetected SARS-CoV-2 infection).

**Methods:** We examined the relation between mortality for COVID-19, mortality for other causes, and ICU admissions with a SARS-CoV-2-infection in adults aged over 60y in France.

**Results:** The number of deaths with principal causes other than COVID-19 in France between July 2021-June 2022 was greater than the corresponding number between July 2020-June 2021 by 20,860 (95% CI (11241,30421)) after adjusting for pre-pandemic trends in mortality (compared to the increase of 3,078 in the number of deaths with a SARS-CoV-2 infection with principal causes other than COVID-19 between the two time periods). During the period of Omicron circulation (Nov. 1, 2021 - Nov. 13, 2022), there was a strong association between the rates of ICU admission with a SARS-CoV-2 infection in adults aged over 60y and (a) rates of COVID-19 deaths (correlation=0.96 (0.92,0.97)); (b) rates of mortality with principal causes other than COVID-19 (correlation=0.89 (0.82,0.94)). Proportions of ICU admissions for causes other than COVID-19 among all ICU admissions with a SARS-CoV-2 infection in older adults were lower during the periods when rates of COVID-19 disease in the community were higher.

**Conclusions:** The significant increase in mortality with principal causes other than COVID-19, as well as the decreases in the proportions of ICU admissions for causes other than COVID-19 among all ICU admissions with a SARS-CoV-2 infection in older adults during the periods when rates of COVID-19 disease in the community were higher suggest under-detection of Omicron infections in associated complications that did not manifest themselves as COVID-19, which is related to the treatment of SARS-CoV-2 infection in those complications.

## Introduction

With the emergence of the Omicron variant of SARS-CoV-2, it became clear that the relative risk for complications, including hospital admission and death in adults is lower for the Omicron variant compared to the Delta variant, though those relative risks vary with age, with the relative risk for severe outcomes, including death for Omicron vs. Delta being greatest for the oldest adults [1,2]. At the same time, appearance of the Omicron variant led to an increase in the proportion of severe outcomes (including hospitalizations and ICU admissions) in SARS-CoV-2-positive patients that were for a cause other than COVID-19 – see for example the data from France on hospitalizations and ICU admissions with SARS-CoV-2 infection for another cause vs. admissions for COVID-19 [3]. This is related to differences in disease manifestation for Omicron infections vs. Delta infections for both emergency department (ED) admissions [4], hospitalizations [5], and admissions to critical care [6]. Those changes also led to changes in patterns of SARS-CoV-2-associated mortality. In France, between Nov. 1, 2021 --July 31, 2022, in addition to 33,353 deaths with a principal cause of COVID-19 [7], there were at least 9,638 deaths with a confirmed SARS-CoV-2 infection with a principal cause other than COVID-19 ([7,8], with some death certificates not being received by CepiDc [8]) – thus, at least 22.4% of all deaths with a confirmed SARS-COV-2 infection during the Omicron period had a principal cause other than COVID-19. For comparison, for the period up to Oct. 31, 2021, there were 115,322 deaths in France with the principal cause of COVID-19, as well as (at least) 11,783 deaths in SARS-CoV-2-positive patients with a principal cause other than COVID-19 [7,8]. Moreover, there were likely additional Omicron-associated deaths in France for which SARS-CoV-2 infection was not detected. In Australia, 21.5% of all deaths with a confirmed SARS-COV-2 infection during the Omicron period had a principal cause other than COVID-19 [9]; among deaths with a confirmed SARS-CoV-2 infection for other principal causes, 26.7% were for cancer, 24.6% were for diseases of the circulatory system, and 20.8% were for dementia [9]. We note that case fatality rates with Omicron-associated COVID-19 disease are relatively high in cancer patients, even the vaccinated ones (Figure 2 in [10]); additionally, while risks of circulatory complications for COVID-19 disease associated with earlier SARS-CoV-2 variants are documented (e.g. [11]), risks of circulatory complications associated with Omicron infections require a better characterization.

In this study we examine mortality with principal causes other than COVID-19 in France during the pandemic, including changes that took place with the emergence of the Omicron variant, as well as mortality with the principal cause of COVID-19, and the relation between ICU admissions with a SARS-CoV-2 infection in adults aged over 60y during the Omicron period and mortality with the principal cause of COVID-19, as well as mortality with other principal causes. We also evaluate temporal changes in the proportion of ICU admissions for causes other than COVID-19 among all ICU admissions with a SARS-CoV-2 infection in older adults. The aim is to understand patterns in mortality with principal causes other than COVID-19, particularly in relation to Omicron circulation in the community, to assess the relation between ICU admissions with a SARS-CoV-2 infection in adults aged over 60y and mortality for different causes, to examine whether there is evidence for under-detection of SARS-CoV-2 infection in associated complications that don’t manifest themselves as COVID-19 disease and lead to fatal outcomes, which is also related to the treatment of those SARS-CoV-2 infections.

## Methods

### Data

Data on the daily number of deaths for all causes in France starting 2015 are available in [12]. Data on the daily number of deaths with the principal cause of COVID-19 are available in [7], whereas data on the daily number of deaths for any cause with confirmed SARS-CoV-2 infection in France are available in [8]. Data on the rates of ICU admissions for COVID-19, as well as for another cause with a confirmed SARS-CoV-2 infection in different age groups in France are available in [3]. Data on the age structure for the French population in 2022 are available in [13], whereas data on the population in France during previous years are available in [14]. Data on the weekly numbers of influenza-like illness (ILI) consultations in metropolitan France are available from the French sentinel surveillance [15]. Sentinel data on testing of respiratory specimens for the different influenza subtypes are available from WHO FluNET [16].

### Temporal trends in mortality prior to the pandemic

To understand temporal trends in mortality prior to the pandemic, we employ a previously developed model [17,18] that expresses weekly mortality rates as a combination of influenza-associated mortality rates, baseline weekly rates for non-influenza mortality (with annual periodicity) and (a linear) temporal trend. The model is calibrated against data for the 2015-2019 period, with the inferred trend in mortality extended to the pandemic period to assess changes in non-COVID-19 mortality during the pandemic period. We note that this model was first applied to the US data [17], and then to data from several countries, including the EU population [19].

Not all influenza-like illness (ILI) consultations in the French sentinel data [15] correspond to influenza infections, and those that do, correspond to infection with different influenza subtypes. For each major influenza subtype (e.g. A/H3N2), we define an indicator for the incidence of that subtype on week *t*(e.g. *A/H3N2(t)*) as

*A/H3N2(t)* = Rate of ILI consultations per 100,000 persons on week *t*in [15] * Percent of respiratory specimens on week *t*in [16] positive for influenza A/H3N2 (1)

We regress weekly mortality rates per 100,000 persons in France between 2015-2019 linearly against the following covariates: incidence indicators for the major influenza subtypes (A/H1N1, A/H3N2, B/Yamagata and B/Victoria), baseline weekly rates for non-influenza mortality (with annual periodicity) and the temporal trend --see [18] for additional details. We use periodic cubic splines to model the mortality rates for the baseline whose shape is unknown [17]. The trend is modelled as a linear polynomial in time (week). Finally, to account for the autocorrelation in the noise, we use a bootstrap procedure (resampling the noise on different weeks) to estimate the confidence bounds for various quantities evaluated in the model [17].

### Mortality for COVID-19 and for other principal causes during the pandemic period and ICU admissions with a SARS-CoV-2 infection in older adults

Using the estimates for the trend in mortality prior to the pandemic in the previous section of the Methods, we assess temporal changes in non-COVID-19 mortality during the pandemic in relation to temporal trends in mortality. We also examine the relation between the weekly rates of ICU admissions with a SARS-CoV-2 infection in adults aged over 60y and (a) weekly rates of deaths for COVID-19; (b) weekly rates of deaths with principal causes other than COVID-19 in France during the Omicron period (Nov. 1, 2021 – Nov. 13, 2022). Based on published data in France [20,21], we consider a two-week lag between ICU admissions and deaths for COVID-19 in the correlation analysis. For deaths with principal causes other than COVID-19, their source is unlikely to be ICU admissions for COVID-19; even for some ICU admissions for other causes with a SARS-CoV-2 infection, the resulting death can be coded as a COVID-19 death. Therefore, we use rates of ICU admissions with a SARS-CoV-2 infection as an indicator (correlate) for SARS-CoV-2-associated mortality rates for non-COVID-19 principal causes of death, with no lag between ICU admission rates and rates of mortality for principal causes other than COVID-19 used in the correlation analysis. Finally, we examine temporal changes in proportions of ICU admissions for causes other than COVID-19 among all ICU admissions with a SARS-CoV-2 infection in different age groups of older adults (60-69y, 70-79y, 80-89y, 90+y), and their relation to the levels of ICU admissions for COVID-19 in the corresponding age groups.

## Results

### Mortality with principal causes other than COVID-19 during the pandemic

Figure 1 plots the weekly rates of mortality for principal causes other than COVID-19 per 100,000 persons in France between week 27, 2020 and week 26, 2022. Those rates have increased during the second half of the study period (week 27, 2021 – week 26, 2022) compared to the first half of the study period. We note that we chose week 26, 2022 to be last week for this analysis because of the high levels of mortality due to a heat wave in July 2022 in France [22]. The number of non-COVID-19 deaths in France between week 27, 2021 – week 26, 2022 was greater than the corresponding number between week 27, 2020 – week 26, 2021 by 20,860 (95% CI (11241,30421)) after adjusting for trends in mortality between 2015-2019 (Methods). At the same time, the number of deaths with a confirmed SARS-CoV-2 infection with a principal cause other than COVID-19 increased by 3,078 between the two time periods [7,8].

**Figure 1:**
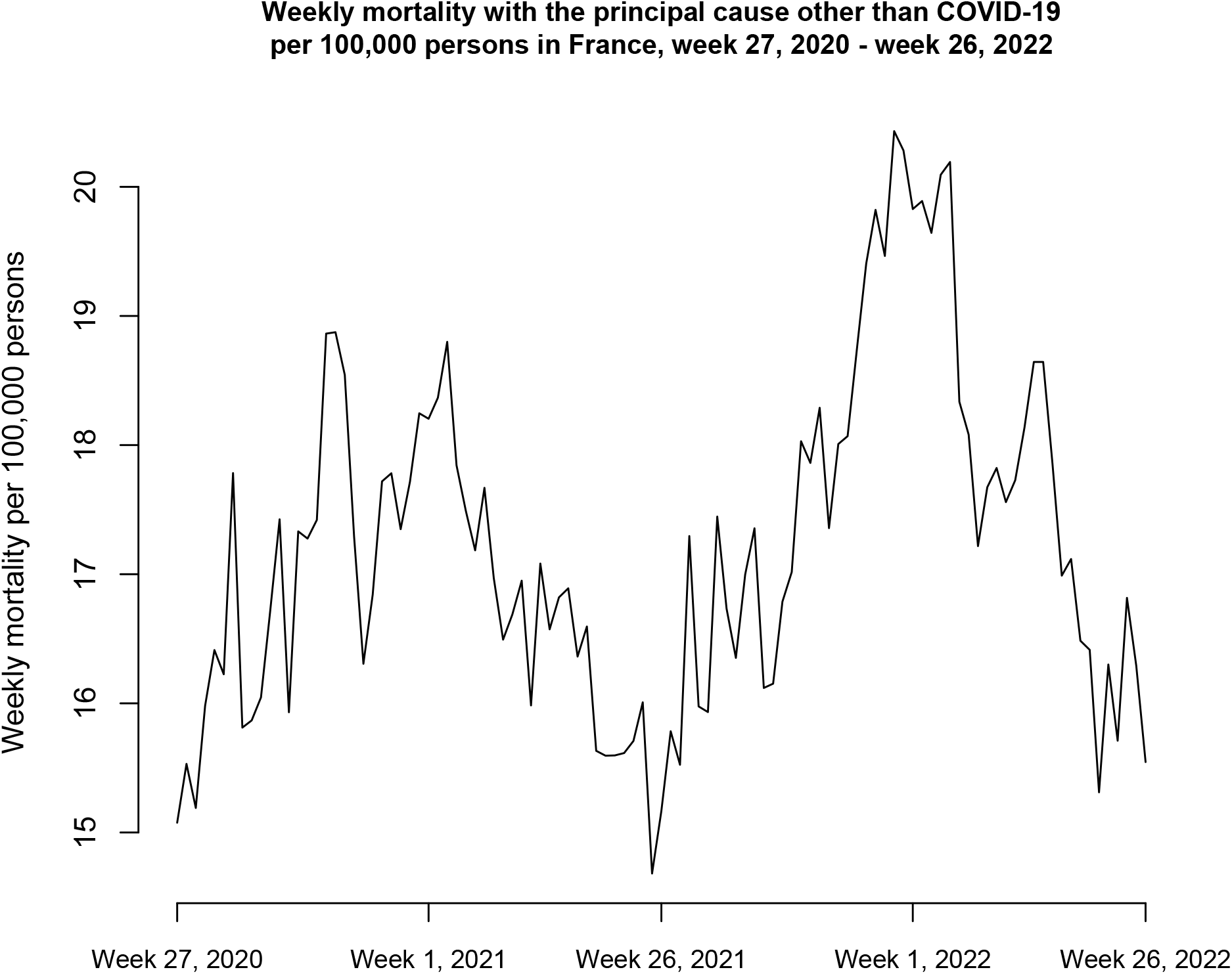
Weekly rates of mortality for a principal cause other than COVID-19 per 100,000 persons in France between week 27, 2020 and week 26, 2022

### COVID-19 mortality, mortality for other principal causes and ICU admissions with a SARS-CoV-2 infection in adults aged over 60y during the Omicron period

Figure 2 (A) suggests a strong temporal relation between the weekly rates of COVID-19 deaths per 100,000 persons in France and rates of ICU admission with a SARS-CoV-2 infection per 100,000 adults aged over 60y two weeks earlier (correlation=0.96 (0.92,0.97) for the period between Nov. 1, 2021 – Nov. 13, 2022). Figure 2 (B) suggests a strong association between the rates of ICU admission with a SARS-CoV-2 infection per 100,000 adults aged over 60y and rates of mortality with principal causes other than COVID-19 per 100,000 persons (correlation=0.89 (0.82,0.94)). We note that the second peak of non-COVID-19 mortality in April, 2022 (Figure 2 (B)) is related not only to SARS-CoV-2 circulation but also to the influenza epidemic in France during that period [23], while the summer peak in mortality in 2022 is related to the heat wave in France during that period [22].

**Figure 2:**
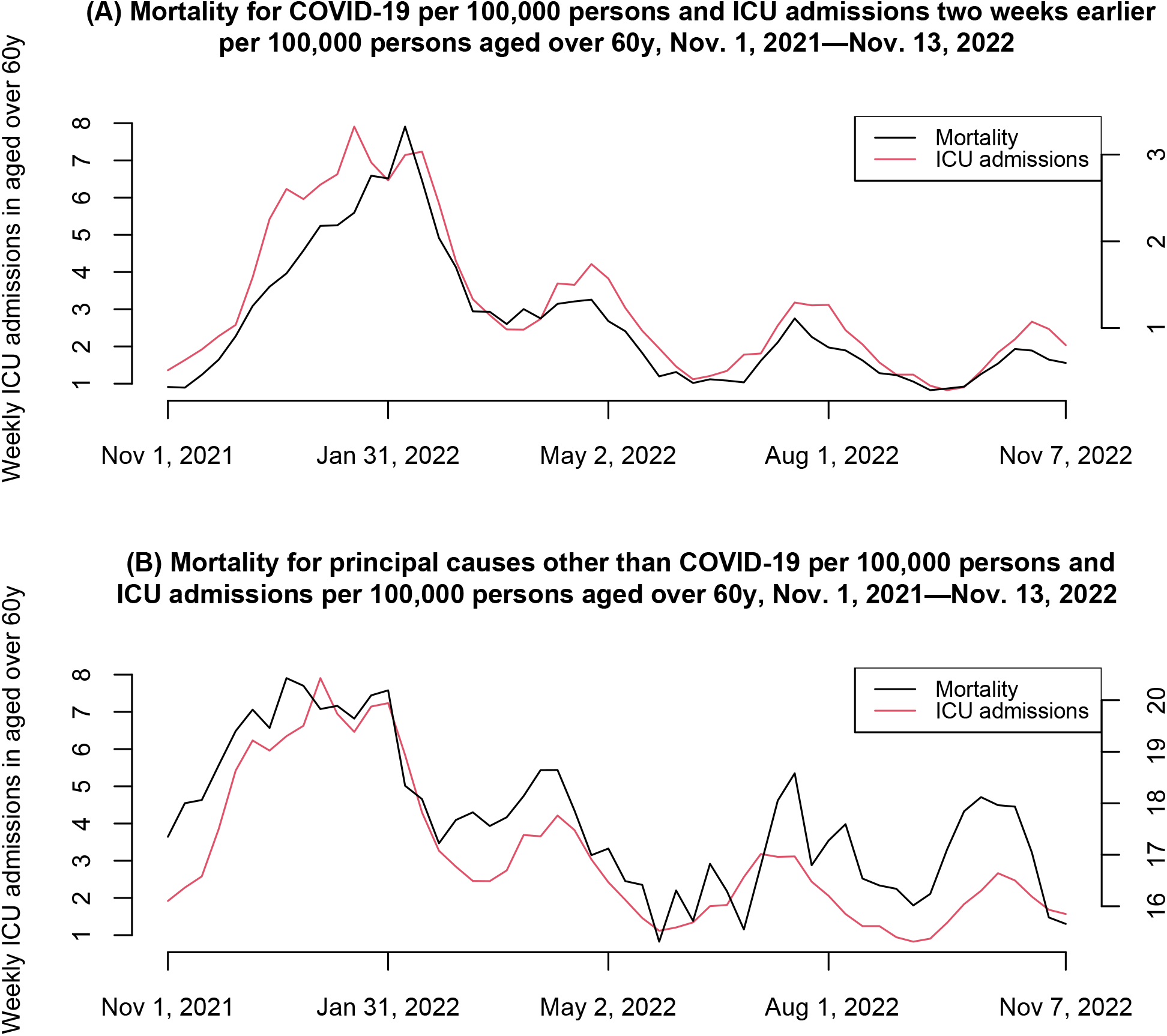
The relation between the weekly rates of ICU admissions with a SARS-CoV-2 infection in adults aged over 60y and (A) weekly rates of deaths for COVID-19; (B) weekly rates of death with principal causes other than COVID-19 in France during the Omicron period (Nov. 1, 2021 – Nov. 13, 2022).

### ICU admissions for COVID-19 and ICU admission for other causes with a SARS-CoV-2 infection in older adults

The proportion of ICU admissions for causes other than COVID-19 among all ICU admissions with a SARS-CoV-2 infection in older adults was lower during the first wave of the Omicron epidemic (thought the end of March, 2022; Figure 3). For the period between March 25, 2022 – Dec. 5, 2022, the proportion of ICU admissions for causes other than COVID-19 among all ICU admissions with a SARS-CoV-2 infection in older adults was lower during periods when COVID-19 disease rates were higher (Figure 3). Correlations between the proportion of ICU admissions for causes other than COVID-19 among all ICU admissions with a SARS-CoV-2 infection and rates of ICU admission for COVID-19 for the period between March 25, 2022 -- Dec. 5, 2022 ranged from -0.4 (−0.5,-0.3) in ages 70-79y to -0.53 (−0.61,-0.44) in ages over 90y (Table 1).

**Table 1:** Correlation between the proportions of ICU admissions for causes other than COVID-19 among all ICU admissions with a SARS-CoV-2 infection and rates of ICU admissions for COVID-19 in different age groups of older adults in France, March 25, 2022 – Dec. 5, 2022

| Age Group | Correlation between the proportions of ICU admissions for causes other than COVID-19 among all ICU admissions with a SARS-CoV-2 infection and rates of ICU admissions for COVID-19, March 25, 2022 – Dec. 5, 2022 |  |
| --- | --- | --- |
|  | Estimate | 95% CI |
| 60-69y | -0.51 | (-0.60, -0.41) |
| 70-79y | -0.40 | (-0.50, -0.30) |
| 80-89y | -0.52 | (-0.60, -0.42) |
| 90+y | -0.53 | (-0.61, -0.44) |

**Figure 3:**
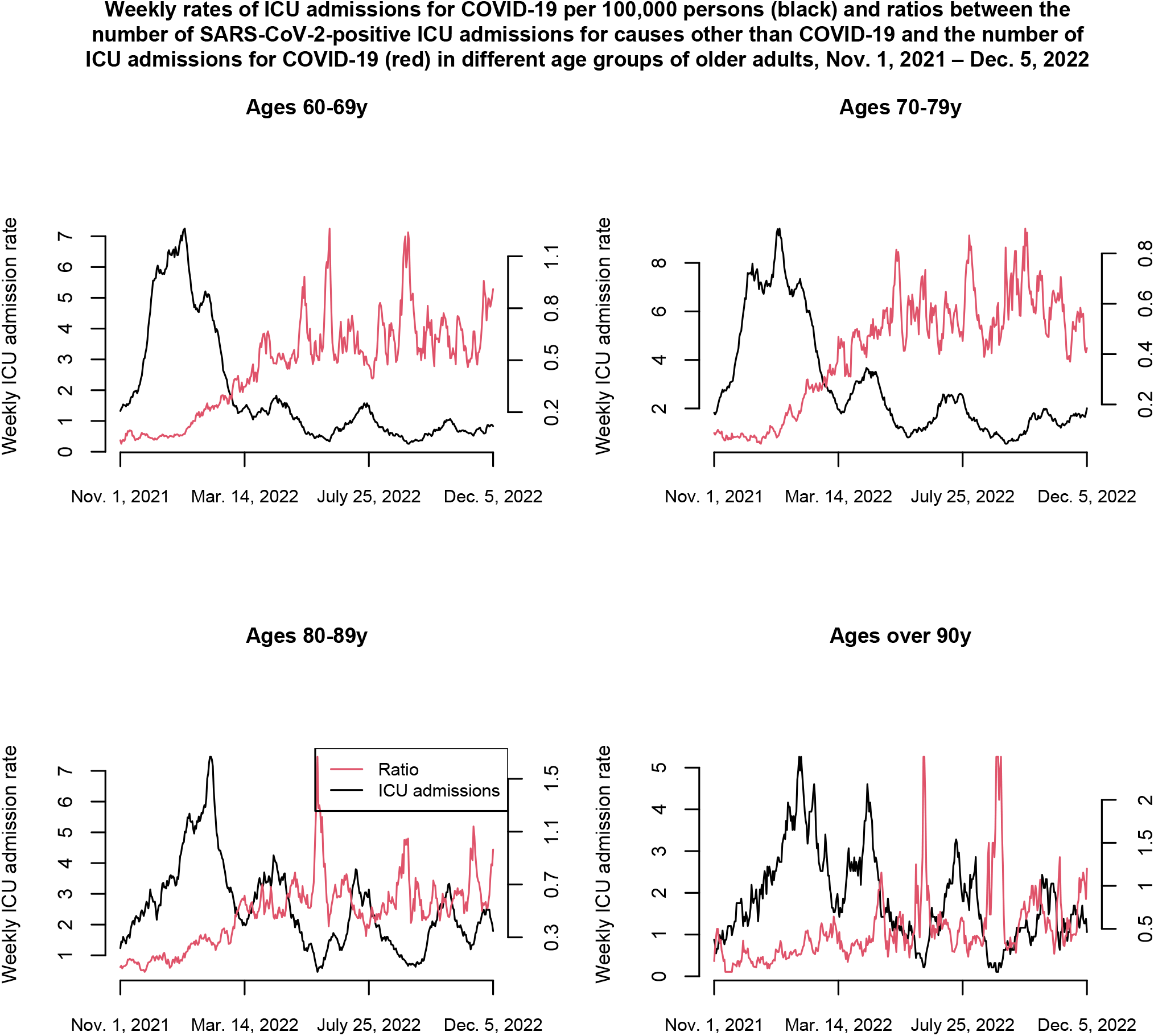
Weekly rates of ICU admissions for COVID-19 per 100,000 persons and ratios between the number of ICU admissions with a SARS-CoV-2 infection for causes other than COVID-19 and the number of ICU admissions for COVID-19 in different age groups of older adults, Nov. 1, 2021 – Dec. 5, 2022

## Discussion

Emergence of the Omicron variant of SARS-COV-2 resulted in changes in the patterns of SARS-CoV-2-related mortality. Case fatality ratios for SARS-CoV-2 infections have decreased significantly compared to past SARS-CoV-2 variants, though less so for the oldest adults [1,2]. At the same time, there was a significant increase in the proportion of deaths with a SARS-CoV-2 infection that were for a principal cause other than COVID-19. For example, in France, between Nov. 1, 2021 and Nov. 13, 2022, at least 22.4% of all deaths with a confirmed SARS-CoV-2 infection were for principal causes other than COVID-19 [7,8]. Additionally, there were Omicron-associated deaths for which SARS-CoV-2 infection wasn’t detected. In this paper, we examined mortality with principal causes other than COVID-19 in France during the pandemic, mortality with the principal cause of COVID-19, and their relation to ICU admissions with a SARS-CoV-2 infection on adults aged over 60y in France. We found a significant increase in mortality with principal causes other than COVID-19 during the later part of the pandemic (with an over 20,000 extra non-COVID-19 deaths between July 2021-June 2022 compared to June 2020-July 2021 after adjusting for pre-pandemic temporal trends in mortality) – at the same time, the number of deaths with a SARS-CoV-2 infection with a principal cause other than COVID-19 increased by 3,078 between the two time periods. Additionally, we found a temporally consistent relation between rates of ICU admissions with a SARS-CoV-2 infection in older adults and mortality for COVID-19 in France, as well as a strong association between rates of ICU admissions with a SARS-CoV-2 infection in older adults and rates of mortality with principal causes other than COVID-19. Finally, we found that proportions of ICU admissions for causes other than COVID-19 among all ICU admissions with a SARS-CoV-2 infection in older adults were lower during periods of active COVID-19 circulation in France, particularly the first wave of the Omicron epidemic, but also during the subsequent epidemic waves when rates of SARS-CoV-2 circulation in the community were high. The significant increase in mortality with principal causes other than COVID-19 during the Omicron period, as well as the decreases in the proportions of ICU admissions for causes other than COVID-19 among all ICU admissions with a SARS-CoV-2 infection in older adults during the periods when rates of COVID-19 disease in the community were higher suggest under-detection of Omicron infections in associated complications that don’t manifest themselves as COVID-19 disease, such as complications related to cancer, circulatory disease, and dementia/Alzheimer’s disease [9], which is related to the treatment of SARS-CoV-2 infection in those complications.

Where are various reasons behind changes in patterns of mortality with principal causes other than COVID-19 (non-COVID-19 mortality) in France during the course of the pandemic. One of those reasons is change in the patterns of circulation of different separatory viruses, including influenza which affect rates of severe outcomes, including deaths [24,25]. In particular, the peak in non-COVID-19 mortality in April 2022 (Figure 2) was affected by the influenza epidemic in France during that period [23] (as well as by the increase in Omicron circulation (Figure 2)). At the same time, the increase of over 20,000 in the number of deaths for causes other than COVID-19 cannot be explained by the increase of 3,078 in the number of deaths with a confirmed SARS-COV-2 infection for causes other than COVID-19, as well as the contribution of influenza to the mortality peak in the Spring of 2022 in France, providing evidence for under-detection of SARS-CoV-2 infections in associated mortality. There can be different reasons behind the temporal changes in the proportion of ICU admissions for] causes other than COVID-19 among all ICU admissions with a SARS-CoV-2 infection in older adults (Figure 3 and Table 1), including long-term changes in disease manifestation for Omicron infections, and changes in coding for the cause of ICU admission. However, the consistent declines in the proportion of ICU admissions for causes other than COVID-19 among all ICU admissions with a SARS-CoV-2 infection in older adults during periods when rates of ICU admission with COVID-19 were higher suggest further evidence for the fact that complications stemming from SARS-CoV-2 infections which don’t manifest themselves clearly as COVID-19 disease are undetected during different stages of the Omicron epidemic.

## Conclusions

With the emergence of the Omicron variant, there was a significant increase in mortality for principal causes other COVID-19. We found an increase of over 20,000 in the number of deaths for principal causes other than COVID-19 between July 2021-June 2022 compared to June 2020-July 2021 after adjusting for pre-pandemic temporal trends in mortality. This increase cannot be explained by a much more modest increase in the number of deaths with a confirmed SARS-CoV-2 infection with principal causes other than COVID-19, as well as changes in the contribution of other respiratory viruses to mortality. Additionally, we found a strong association between the rates of ICU admission with a SARS-CoV-2 infection in adults aged over 60y and rates of non-COVID-19 mortality during the Omicron epidemic in France, as well as the decline in the proportion of ICU admissions for causes other than COVID-19 among all ICU admissions with a SARS-CoV-2 infection in older adults during periods when rates of COVID-19 disease in the community were higher. All of this suggests a substantial contribution of Omicron infections to mortality for principal causes other than COVID-19 (such as cancer, circulatory disease, and dementia/Alzheimer’s disease [9]), as well as under-detection of Omicron infections in associated complications that don’t manifest themselves as COVID-19 disease, which might be related to the treatment of SARS-CoV-2 infection in those complications.

## Data Availability

This study is based on aggregate, de-identified data available through refs. 3,7,8,12-16.

https://www.santepubliquefrance.fr/dossiers/coronavirus-covid-19/coronavirus-chiffres-cles-et-evolution-de-la-covid-19-en-france-et-dans-le-monde

https://covid19.who.int/

https://opendata.idf.inserm.fr/cepidc/covid-19/telechargements

https://www.insee.fr/fr/statistiques/4931039?sommaire=4487854#tableau-figure1

https://www.insee.fr/fr/statistiques/2381472#graphique-Donnes

https://www.insee.fr/fr/statistiques?debut=0&theme=1

https://app.powerbi.com/view?r=eyJrIjoiNjViM2Y4NjktMjJmMC00Y2NjLWFmOWQtODQ0NjZkNWM1YzNmIiwidCI6ImY2MTBjMGI3LWJkMjQtNGIzOS04MTBiLTNkYzI4MGFmYjU5MCIsImMiOjh9

